# The prostate tissue-based telomere biomarker as a prognostic tool for metastasis and death from prostate cancer after prostatectomy

**DOI:** 10.1101/2021.12.01.21267154

**Authors:** Christopher M. Heaphy, Corinne E. Joshu, John R. Barber, Christine Davis, Jiayun Lu, Reza Zarinshenas, Edward Giovannucci, Lorelei A. Mucci, Meir J. Stampfer, Misop Han, Angelo M. De Marzo, Tamara L. Lotan, Elizabeth A. Platz, Alan K. Meeker

## Abstract

**Purpose:** Current biomarkers are inadequate prognostic predictors in localized prostate cancer making treatment decision-making challenging. Previously, we observed that the combination of more variable telomere length among prostate cancer cells and shorter telomere length in prostate cancer-associated stromal cells – the telomere biomarker – is strongly associated with progression to metastasis and prostate cancer death after prostatectomy independent of currently used pathologic indicators.

**Experimental Design:** We optimized our method allowing for semi-automated telomere length determination in single cells in fixed tissue, and tested the telomere biomarker in tissue microarrays from five cohort studies of men surgically treated for clinically localized disease (N=2,255). We estimated the relative risk (RR) of progression to metastasis (N=311) and prostate cancer death (N=85) using models appropriate to each study’s design adjusting for age, prostatectomy stage, and tumor grade, which then we meta-analyzed using inverse variance weights.

**Results:** Compared with men who had less variable telomere length among prostate cancer cells and longer telomere length in prostate cancer-associated stromal cells, men with the combination of more variable and shorter telomere length, had 3.76-times the risk of prostate cancer death (95% CI 1.37-10.3; p=0.01) and had 2.23-times the risk of progression to metastasis (95% CI 0.99-5.02, P=0.05). The telomere biomarker was associated with prostate cancer death in men with intermediate risk disease (Grade Groups 2/3: RR=9.18, 95% CI 1.14- 74.0, p=0.037) and with PTEN intact tumors (RR=6.74, 95% CI 1.46-37.6, p=0.015).

**Conclusions:** The telomere biomarker is robust and associated with poor outcome independent of current pathologic indicators in surgically-treated men.

**Translational Relevance:** Current prognostic biomarkers in localized prostate cancer are inadequate imperfect predictors; therefore, new biomarkers are needed to improve the prognostic classification and management of these patients. In a five-cohort study, we confirmed that the tissue-based telomere biomarker – the combination of more variable telomere length among prostate cancer cells and shorter telomere length in prostate cancer-associated stromal cells – was associated with progression to metastasis and prostate cancer death independent of currently used prognostic indicators after prostatectomy for clinically-localized disease. Importantly, the telomere biomarker was associated with poor outcome in men with intermediate risk disease, as well as in men with intact PTEN tumors. Thus, this tissue-based telomere biomarker has the translational potential to improve treatment and surveillance decision-making.

## INTRODUCTION

Currently used pathologic prognostic indicators do not adequately predict prostate cancer behavior in men with clinically-localized disease (1). To target men with appropriate, individualized treatment strategies or surveillance, new molecular markers that improve prognostic accuracy beyond the currently used pathologic stage and Gleason sum (or Grade Group) are urgently needed. One such molecular tissue-based marker is the measurement of telomeres - the repetitive DNA sequence at the ends of the chromosomes, which are pivotal for maintenance of genome integrity (2–4). Telomere dysfunction is common in precancerous lesions (e.g., high-grade prostatic intraepithelial neoplasia) and continued critical telomere shortening and chromosomal breakage-fusion-bridge cycles leads to chromosomal instability, thereby driving malignant transformation and cancer progression (5, 6).

In our prior cohort study, we discovered that men surgically treated for clinically localized disease who had more variable telomere length among cancer cells and shorter telomere length in prostate cancer-associated stromal cells (i.e., the “telomere biomarker”) had a substantially higher risk of progression to metastasis and prostate cancer death than men who had less variable telomere length in cancer cells and longer telomere length in cancer-associated stromal cells (7). Importantly, these findings were independent of the pathologic prognostic indicators and added prognostic information to those indicators including in men with Gleason 7 disease, who tend to have to have a more variable course. Notably, men with the less variable/longer combination of the telomere biomarker rarely died of their cancer over 15 years. The telomere biomarker was not prognostic for death from other causes, supporting its specificity for aggressive disease. Moreover, we found that variability in telomere length among cancer cells, one component of the telomere biomarker, was associated with recurrence after surgery. Our prior work used the manual method that we developed to measure telomere length with single cell resolution, assessing 30-50 user-selected cells per cell type, while maintaining tissue architecture in archival, formalin-fixed paraffin-embedded (FFPE) prostate tissues (5).

Since our manual method was very labor intensive and possibly susceptible to user bias during the manual selection of the cells to be analyzed, we now have developed a robust, semi-automated method to quantitate cell type-specific telomere length at single cell resolution. Our semi-automated method is based on performing telomere-specific FISH combined with multiplex immunofluorescence to detect a basal cell-specific cytokeratin, prostate epithelial-cell specific nuclear markers (NKX3.1 and FOXA1), and lymphocyte-specific markers (CD3 and CD20) using FFPE tissue samples. This staining process is then followed by semi-automated slide scanning and multi-channel acquisition of fluorescent microscopy images. Cell-type specific telomere and nuclear DNA content data are then obtained from these collected images via semi-automated image analysis allowing us to measure telomere length in all cells of the specified type in focal plane without selection by the operator.

While our original findings pointed to the potential prognostic utility of the telomere biomarker, we next sought to confirm, in a larger study of five cohorts, that the telomere biomarker indeed is independently associated with risk of poor outcome in men surgically treated for clinically-localized prostate cancer using our optimized, semi-automated method. We expanded the number of men surgically treated for prostate cancer from 596 to 2,255, number of metastatic or rapidly rising PSA events from 54 to 311, and number of prostate cancer deaths from 46 to 85. We again assessed the telomere biomarker in men with Gleason 7 disease (Grade Groups 2/3 (8)), a group for whom clinical management decisions are challenging. In addition, we assessed, for the first time, whether the telomere biomarker is associated with outcomes among men with PTEN intact tumors, as PTEN loss has been associated with poor prognosis (9–11).

We confirm here that the telomere biomarker is independently associated with progression to metastasis and prostate cancer death in men surgically treated for prostate cancer, including in men with intermediate disease (Grade Groups 2/3) and in men with PTEN intact tumors. We also confirm that variability in telomere length among cancer cells, but not the telomere biomarker, is associated with recurrence. Thus, the telomere biomarker has the promise to aid in better treatment and surveillance decision-making for these men.

## METHODS

### Study Populations and Designs

We used tissue and data from five cohorts: Health Professionals Follow-up Study (HPFS), the cohort in which we originally described the telomere biomarker (7), Physicians’ Health Study (PHS), Johns Hopkins Recurrence Nested Case-Control Study, and two Johns Hopkins Intermediate-High-Risk Case-Cohort studies. This work was approved by the IRB at the Johns Hopkins University. The cohort study protocol was approved by the institutional review boards of the Brigham and Women’s Hospital and Harvard T.H. Chan School of Public Health, and those of participating registries as required. The study populations were used as designed and are described below and further in **Supplemental Methods**.

#### Health Professionals Follow-up Study (HPFS)

Investigators at Harvard previously developed a cohort study of prostate cancer cases as described (12); prostatectomy tissue from these men were arrayed on 5 TMAs. The HPFS investigators extended follow-up of the men on the original 5 HPFS TMAs (596 of the 631 men were included in the original telomere biomarker paper after exclusions (7)) and added 2 newly constructed TMAs (159 men) to achieve 755 men surgically treated for clinically organ-confined prostate cancer in the analysis. Of these men 227 recurred. Of these, 30 experienced distant metastases, and 68 (46 from original TMAs) men died of their prostate cancer (**Table 1**). HPFS pathologists re-reviewed H&E-stained tissue sections containing prostate cancer and assigned a standardized Gleason sum (13), which we used in the analyses. **Supplement Table 1** shows characteristics of the men in the HPFS who were included in the telomere biomarker analysis. PTEN was previously measured by immunohistochemistry (IHC) (14).

**Table 1.** Number of men who experienced prostate cancer outcomes, number of men in the cohort or comparison group, median time to event, and total follow-up time, 5 cohorts of men surgically treated for clinically localized disease

|  | HPFS | PHS | Johns Hopkins<br>Recurrence | Johns Hopkins<br>Intermediate-High Risk - I | Johns Hopkins<br>Intermediate-High Risk - II |
| --- | --- | --- | --- | --- | --- |
| Study design | Cohort | Cohort | Nested case-control | Case-cohort | Case-cohort |
| Prostate cancer outcome |  |  |  |  |  |
| Recurrence<sup>1</sup> | 227 | 51 | 376 | - | - |
| Metastasis | 30 | 5 | - | 115 | 161<sup>3</sup> |
| Prostate cancer death | 68<sup>2</sup> | 17 | - | - | - |
| Cohort/controls/subcohort | 755<sup>2</sup> | 151 | 376 | 253 | 192 |
| Total number of men evaluated | 755 | 151 | 752 | 306<sup>4</sup> | 291<sup>5</sup> |
| Median event and follow-up times (years) |  |  |  |  |  |
| Case time to recurrence | 3.8 | 4.2 | 2.0 | - | 1.0 |
| Case time to metastasis | 9.7 | 8.8 |  | 4.0 | 3.0 |
| Case time to prostate cancer death | 10.0 | 5.5 | - | - | - |
| Cohort/control follow-up | 14.6 | 13.8 | 6.0 | 10.0 | 3.0 |

HPFS – Health Professionals Follow-up Study, PHS – Physicians’ Health Study
<sup>1</sup>Includes biochemical recurrence.
<sup>2</sup>Of these, 50 prostate cancer deaths and 596 men were included in the original study in which we described the telomere biomarker.
<sup>3</sup>Of these, 125 had rapidly rising PSA (doubling time <10 months) indicative of a high risk of metastasis.
<sup>4</sup>62 men in the subcohort progressed and 53 men outside of the subcohort progressed (253 subcohort + 53 progressed outside of the subcohort = 306).
<sup>5</sup>62 men in the subcohort progressed and 99 men outside of the subcohort progressed (192 subcohort + 99 progressed outside of the subcohort = 291).

#### Physicians’ Health Study (PHS)

Investigators at Harvard University previously developed a cohort study of men diagnosed with prostate cancer in the PHS between 1983 and 2009; prostatectomy tissue from these men were arrayed on 5 TMAs. Of these men, 45 developed distant metastases or died of their prostate cancer over a mean of 12.9 years of follow-up (15). This PHS sub-study was designed to identify tissue-based prognostic markers for lethal prostate cancer (12,15,16). Two study pathologists re-reviewed all specimens to provide a standardized Gleason scoring, which was previously documented to improve prediction of death from prostate cancer (13). PTEN was previously measured by IHC (14).

In the first run for this telomere biomarker study, less than half of the TMA spots were useable across the 5 TMAs. The PHS investigators subsequently provided sections from replica TMAs, which were available on 4 of the 5 TMAs; again, less than half of the spots were useable. Reasons included some the spots no longer contained tumor because the tissue fell out of the section matrix or was missing initially, or had high auto-fluorescent background (depending on the TMA, ∼15-20% of spots) that prohibited determination of telomere length. Thus, we included in the analysis 151 men with evaluable spots in the analysis. Of these men 51 recurred, 5 experienced distant metastases, and 17 men died of their prostate cancer (**Table 1**).

**Supplement Table 2** show characteristics of men in the PHS who were included in the telomere biomarker analysis.

#### Johns Hopkins Recurrence Nested Case-Control Study

At the Brady Urological Institute at Johns Hopkins, we previously developed a Recurrence Nested Case-Control Study and have used it to investigate tissue prognostic markers, such as PTEN by IHC (17) and intratumoral mast cells (18). Cases and controls were drawn from men who had had a prostatectomy for clinically localized prostate cancer at Johns Hopkins between 1993 and 2001 and had not had hormonal or radiation therapy before surgery or adjuvant therapy before recurrence. Cases (N=524) were men with biochemical recurrence (re-elevation of PSA ≥0.2 ng/mL), metastasis, or prostate cancer death after surgery by 2004, whichever came first. For each case, a control (N=524) was selected who had not recurred by the case’s date and was matched on age, race, pathological stage, and Gleason sum. 16 TMAs were constructed with matched case-control pairs spotted on the same TMA. Evaluable spots were available for 376 matched recurrence pairs (**Table 1**). **Supplement Table 3** shows case and control characteristics for men included in the telomere biomarker analysis.

#### Intermediate-High Risk Case-Cohort Study – I

The Brady Urological Institute at Johns Hopkins developed an intermediate and high-risk case-cohort study and associated 9 TMAs for use in identifying tissue markers of metastasis in men at higher risk for poor outcome (19–21). Men were selected from the Johns Hopkins radical prostatectomy (RP) clinical research database (>20,000 patients, of which >13,000 have long term follow-up) of men who underwent RP at Johns Hopkins between 1992 and 2010 and who had intermediate or high-risk disease by the Cancer of the Prostate Risk Assessment (CAPRA)-S score ≥3 (22). Men with metastatic disease or positive lymph nodes detected by imaging before surgery, men who received neoadjuvant therapy, and men who did not have a PSA nadir of <0.2 ng/mL post-surgery were excluded. Men who received hormone, chemo-, or radiation therapy after surgery but before detection of metastasis by imaging were excluded to be able to address how biomarkers are associated with metastatic outcome without the interference of treatment during follow-up. The eligible cohort consisted of 745 men from which a ∼35% sample was randomly selected. Men who did not develop metastases in the subcohort and in the eligible cohort were shown to be similar (19). The study includes 267 men in the subcohort and 119 men (including 64 in the subcohort) who progressed to metastasis. PTEN was previously measured by IHC (23).

Included in the telomere analysis were 253 men from the subcohort (94.8%) and 115 of the metastatic cases (96.6%) (**Table 1**). **Supplement Table 4** shows characteristics of all of the men, men in the subcohort, and cases outside of the subcohort who were included in the telomere biomarker analysis.

#### Intermediate-High Risk Case-Cohort Study – II

The Brady Urological Institute subsequently developed a second case-cohort study of men with intermediate and high-risk prostate cancer and associated 9 TMAs also for use in identifying tissue markers of progression to disease with a lethal phenotype in men at higher-risk. Men with intermediate and high-risk disease who underwent RP between 2007 and 2015, had not had neoadjuvant therapy, and had data available on all required clinical and outcome variables in the Johns Hopkins radical prostatectomy database were selected. Risk was based on the based on the D’Amico classification at the time of biopsy (24): intermediate - stage T2b or Gleason 7 or PSA >10 and ≤20 ng/mL; and high -stage T2c or PSA >20 ng/mL or Gleason ≥8). This left 3,762 eligible men in the source population, in whom 204 experienced metastases or rapidly rising PSA (PSA doubling time <10 months). For feasibility, 121 of the cases were sampled. Then, a random subcohort (N=254, 6.8% of 3,762) of size of double the number of cases was sampled. Of the 121 cases, 19 occurred in the subcohort. Median follow-up of the subcohort was 3.0 years.

Median time to lethal progression in the cases was 3.5 years. Men who received chemo-, radiation, or hormone therapy between surgery and detection of metastasis were not excluded; any treatment subsequent to prostatectomy cannot affect the telomere biomarker in the primary tumor. **Supplement Table 5** shows characteristics of all of the men, men in the subcohort, and cases outside of the subcohort who were included in the telomere biomarker analysis.

### Measurement of Telomere Length

For the original HPFS TMAs (N=5) we used the non-automated method described in Heaphy et al. (7). For the additional HPFS TMAs (N=2), and the TMAs from the PHS, Johns Hopkins Recurrence Nested Case-Control Study, and the Johns Hopkins Intermediate-High Risk Case-Cohort Study I and II, we used our semi-automated, optimized method described in Heaphy et al. (25) and summarized here.

#### Telomere-specific FISH and Immunostaining

Deparaffinized TMA slides were stained for telomeres by telomere-specific FISH and co-labeled by multiplex immunofluorescence (see **Supplemental Methods**).

#### Microscopy and Image Analysis

The TissueFAXS Plus (Tissue Gnostics, Vienna, Austria) automated microscopy workstation, which contains an 8-slide ultra-precise motorized stage and utilizes a Zeiss Z2 Axioimager microscope, was used for automated image acquisition. First, a DAPI preview image is captured with a 10X objective to allow for appropriate orientation. Next, the TMA spots are identified and images are captured with a 40X oil objective using the DAPI, GFP, Cy3, and Cy5 filters. An autofocus algorithm in the DAPI filter and the extended focus parameter by capturing 3 steps above and below (step size=0.8 µm) was utilized. An entire TMA with 400 spots can be imaged in ∼14 hours, which is faster than other current imaging modalities (26). For image analysis, a separate high-performance workstation with the TissueQuest software module to analyze the fluorescent images with precise nuclear segmentation was used (25, 27). A region of interest is set (e.g., stroma or cancer) and processed for nuclear segmentation. If required, exclusion regions were set to exclude benign prostate glands.

#### Categorizing the telomere biomarker

After exporting the data to Excel spreadsheets, we converted them to a SAS dataset and merged the data with the TMA spot individual identifiers. For each man and for each cell, we calculated the ratio of Cy3 dot sum intensity and DAPI dot sum intensity and multiplied by 1000; this ratio is the telomere ratio. The telomere ratio for each nucleus compensates for differences in nuclear cutting planes and ploidy. Separately, for each cohort we performed the following: For each man, we calculated median telomere ratio for cancer-associated stromal cells; this is the first of the two components of the telomere biomarker. For each man, we calculated the standard deviation of telomere ratio for cancer cells; this is second of the two components of the telomere biomarker. We determined whether the man’s median telomere ratio for cancer-associated stromal cells was below the 66^th^ percentile among the men in each TMA set; we categorized this group as having shorter telomeres in cancer-associated stromal cells. We determined whether a man’s standard deviation of telomere ratio for cancer cells was above the 66^th^ percentile of the distribution of the standard deviation of telomere ratio among men in each TMA set; we categorized this group as having more variable telomeres in cancer cells. Based on our prior study (7), individuals who have the combination of shorter telomeres in cancer-associated stromal cells and more variable telomeres in cancer cells have the poorest prognosis; individuals who either have shorter telomeres in cancer-associated stromal cells OR more variable telomeres in cancer cells have an intermediate prognosis; and individuals who have neither shorter telomeres in cancer-associated stromal cells nor more variable telomeres in cancer cells have the best prognosis.

### Statistical Analysis

For all analyses, we used SAS v. 9.4 (Cary, NC). For the HPFS and PHS studies, we used Cox proportional hazards regression to estimate hazard ratios (HRs) and 95% confidence intervals (CI) and adjusted for age at diagnosis, year of surgery, prostatectomy Gleason sum, pathologic stage, and pre-operative PSA. For the Johns Hopkins Intermediate-High Risk Study I and II, we used Cox proportional hazards regression with robust variance correction to estimate HRs and 95% CIs and adjusted for age, race, pathologic stage, prostatectomy Gleason sum, year of surgery, pre-operative PSA (if missing, a separate indicator variable was used), and surgical margins. For the Johns Hopkins Recurrence Nested Case-Control Study, we used conditional logistic regression to estimate odds ratios (ORs, as unbiased estimates of the HR) and 95% CIs and adjusted for year of surgery, primary and secondary Gleason pattern, pre-operative PSA, and surgical margins (cases and controls were matched on age, race, categories of prostatectomy Gleason sum, and categories pathologic stage).

For each cohort, we estimated the association of more (vs. less) variability in telomere length among cancer cells and shorter (vs. longer) telomere length in cancer-associated stromal cells with recurrence, progression to metastasis, and prostate cancer death. Next, for each study we estimated the association between the telomere biomarker and these same outcomes using the less variable/longer combination as the reference group. We repeated this analysis stratified by Gleason sum (<7, 7, >7) and by PTEN status (intact, null).

Because of the differences in study design, disease severity at diagnosis, timing relative to the PSA era, the telomere length determination method used (original 5 HPFS TMAs non-automatic, all others optimized semi-automated), the fact that the TMAs were run in different batches with slight modifications to the scanning parameters, we could not pool the data from the five cohorts. Instead, we used a meta-analytic approach to obtain summary estimates. To do so, we used inverse variance weights.

## RESULTS

### Characteristics of the men in the 5 cohorts

We included men from five studies who were treated by radical prostatectomy for clinically-localized prostate cancer. Characteristics of each cohort are shown in **Supplemental Tables 1-5**. **Table 1** provides the numbers of men and events included from each study. In total, we included 1,659 newly studied men along with 596 men we previously studied for whom we extended their follow-up. Across these studies, 654 men experienced recurrence, 311 men progressed to distant metastasis or had rapidly rising PSA indicative of likely metastasis (PSA doubling time <10 months), and 85 men died of their prostate cancer. The median follow-up times ranged from 3.0 to 14.6 years.

### Measurement of telomere length in individual cells of specified type using the newly developed and optimized combined telomere FISH and IF staining method

As we previously demonstrated, telomere-specific FISH signal intensities are linearly proportional to telomere length and can be quantified via digital image analysis (28). In our new method, we have combined telomere-specific FISH with multiplex IF staining to better identify and more easily restrict analysis to specific cell types of interest. As shown in **Figure 1A**, basal-specific cytokeratin positivity (magenta) delineates benign prostate glands, the prostate epithelial-cell specific nuclear markers (NKX3.1 and FOXA1; green) highlight prostatic epithelial cells, and lymphocyte-specific markers (CD3 and CD20; magenta) identify lymphocytes in the surrounding microenvironment. Shown in **Figure 1B** are the telomere FISH signals that are robust in the benign gland, while diminished in the cancer cells. Using nuclear segmentation, the cancer cells (green^+^/magenta^-^) and cancer-associated stromal cells (green^-^/magenta^+^) can be identified; whereas the benign prostatic glands and lymphocytes are excluded from the analysis.

**Figure 1.**
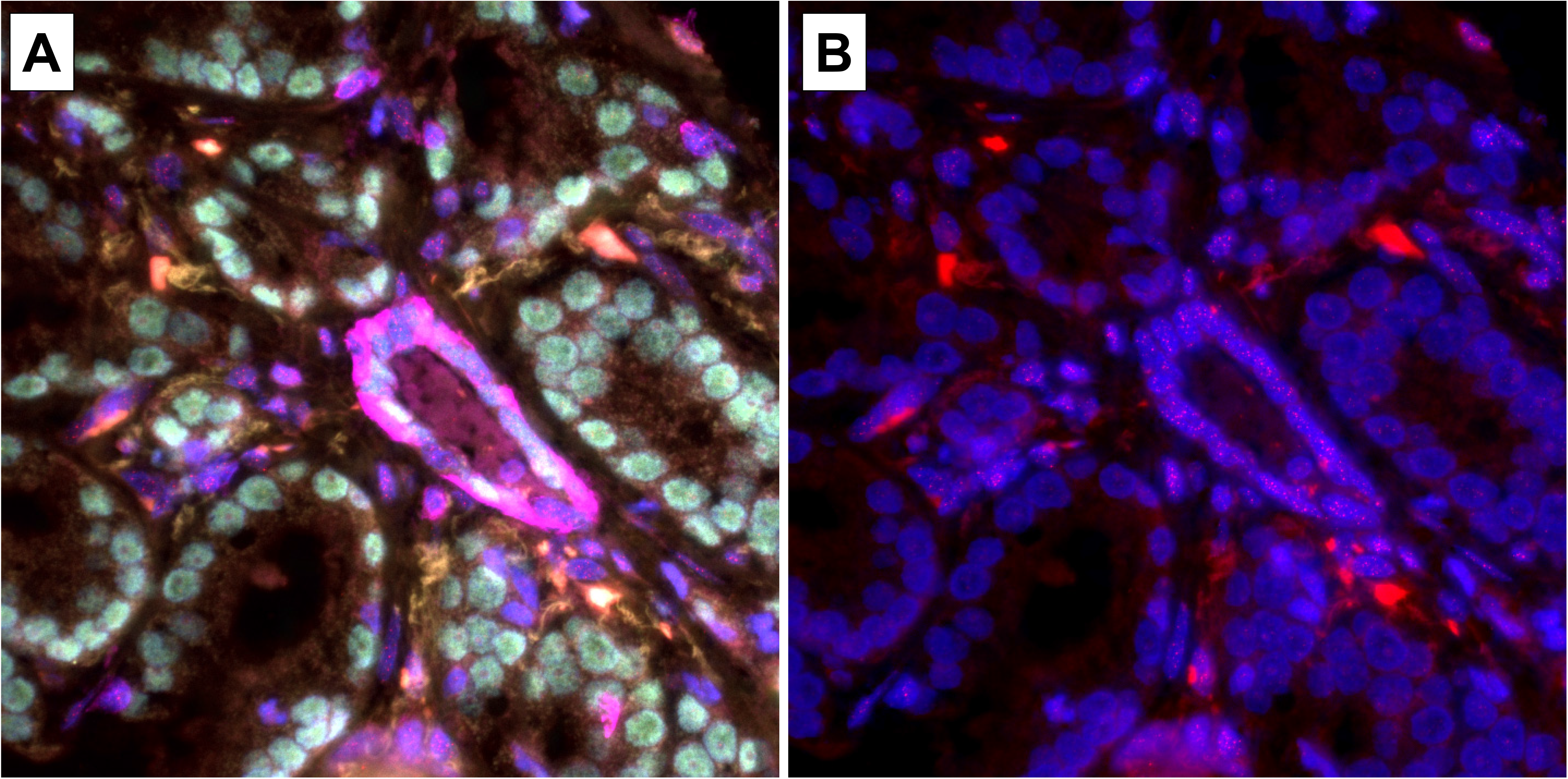
Combined telomere-specific FISH and multiplex immunofluorescence staining in prostate cancer. To measure telomere lengths in individual cells of specific cell type, this newly developed assay is utilized. **A**) In a prostate cancer that contains both benign and cancer regions, a basal-specific cytokeratin (magenta) delineates the benign prostate glands, two epithelial-cell specific nuclear markers (NKX3.1 and FOXA1; green) highlight prostatic epithelial cells, and two lymphocyte-specific markers (CD3 and CD20; magenta) identify lymphocytes in the surrounding tumor microenvironment. **B**) In the same region, the telomeres are highlighted with a Cy3-labeled telomere-specific peptide nucleic acid probe (red). In both images, the DNA is stained with DAPI (blue). Original magnification x400.

### Components of the telomere biomarker and prostate cancer outcomes

**Table 2** shows the summary associations between the two components of the telomere biomarker – variability in telomere length among cancer cells and telomere length in stromal cells – and recurrence, progression to metastasis, and prostate cancer death meta-analyzed across the 5 studies. Considering currently used prognostic factors, more variable telomere length in cancer cells appeared to be similarly associated with a higher risk of each outcome albeit only statistically significant for recurrence. Shorter telomeres in stromal cells were associated with a higher risk of prostate cancer death only (hazard ratio (HR)=1.84, 95% confidence interval (CI) 1.06-3.21, P=0.03). **Supplement Table 6** shows study-specific HRs, and **Supplement Table 7** shows the summary associations excluding the original HPFS study data.

**Table 2.** Meta-analytic summary relative risks (RR) and 95% confidence intervals (CI) of more variable telomere length among prostate cancer cells and shorter telomere length in prostate cancer-associated stromal cells with risk of recurrence, metastasis, and prostate cancer death after prostatectomy for clinically localized disease in 5 cohorts^1^

|  | Number of contributing cohorts | Summary RR<sup>2</sup>, 95% CI |  |
| --- | --- | --- | --- |
|  |  | More variable<sup>3</sup> telomere length among prostate cancer cells (vs less variable) | Shorter<sup>4</sup> telomere length in cancer-associated stromal cells (vs longer) |
| Recurrence | 3 | 1.40<br>1.13-1.74<br>P=0.002 | 0.94<br>0.75-1.17<br>P=0.57 |
| Metastasis | 4<sup>5</sup> | 1.36<br>0.90-2.04<br>P=0.14 | 1.19<br>0.75-1.88<br>P=0.47 |
| Prostate cancer death | 2 | 1.37<br>0.85-2.20<br>P=0.20 | 1.84<br>1.06-3.21<br>P=0.03 |

Bold denotes statistically significant at p<0.05 for a 2-sided test.
<sup>1</sup>50 prostate cancer deaths and 596 men were included in the original study in which we described the telomere biomarker.
<sup>2</sup>Each contributing RR and 95% CI was adjusted for currently used prognostic pathologic factors. RRs were summarized using inverse variance weights.
<sup>3</sup>More variable was defined as the top tertile of variability in telomere length among prostate cancer cells. Less variable was defined as the bottom and middle tertiles.
<sup>4</sup>Shorter was defined as the shortest and middle tertiles of the median telomere length in cancer-associated stromal cells. Longer was defined as the longest tertile.
<sup>5</sup>Includes all events (N=161) from the Johns Hopkins Intermediate-High Risk – II. Including only the 35 metastatic events: more variable length among cancer cells HR=1.39, 95% CI 0.93-2.06 (P=0.11), shorter length among stromal cells HR=1.12, 95% CI 0.71-1.75 (P=0.64).

### The telomere biomarker and prostate cancer outcomes

**Table 3** and **Figure 2** show the summary associations between the telomere biomarker and recurrence, progression to metastasis, and prostate cancer death meta-analyzed across the 5 cohorts. Taking into account the pathologic prognostic markers, compared with men with less variable telomere length in their cancer cells and longer telomere length in their stromal cells, men with more variable telomere length in their cancer cells and shorter telomere length in their stromal cells had 3.76 times the risk of prostate cancer death (95% CI 1.37-10.3, P=0.01) and had 2.23 times the risk of metastasis (95% CI 0.99-5.02, P=0.05); men with the two other combinations had intermediate HRs. In contrast, only the telomere biomarker categories that include more variable telomere length were associated with an increased risk of recurrence.

**Figure 2.**
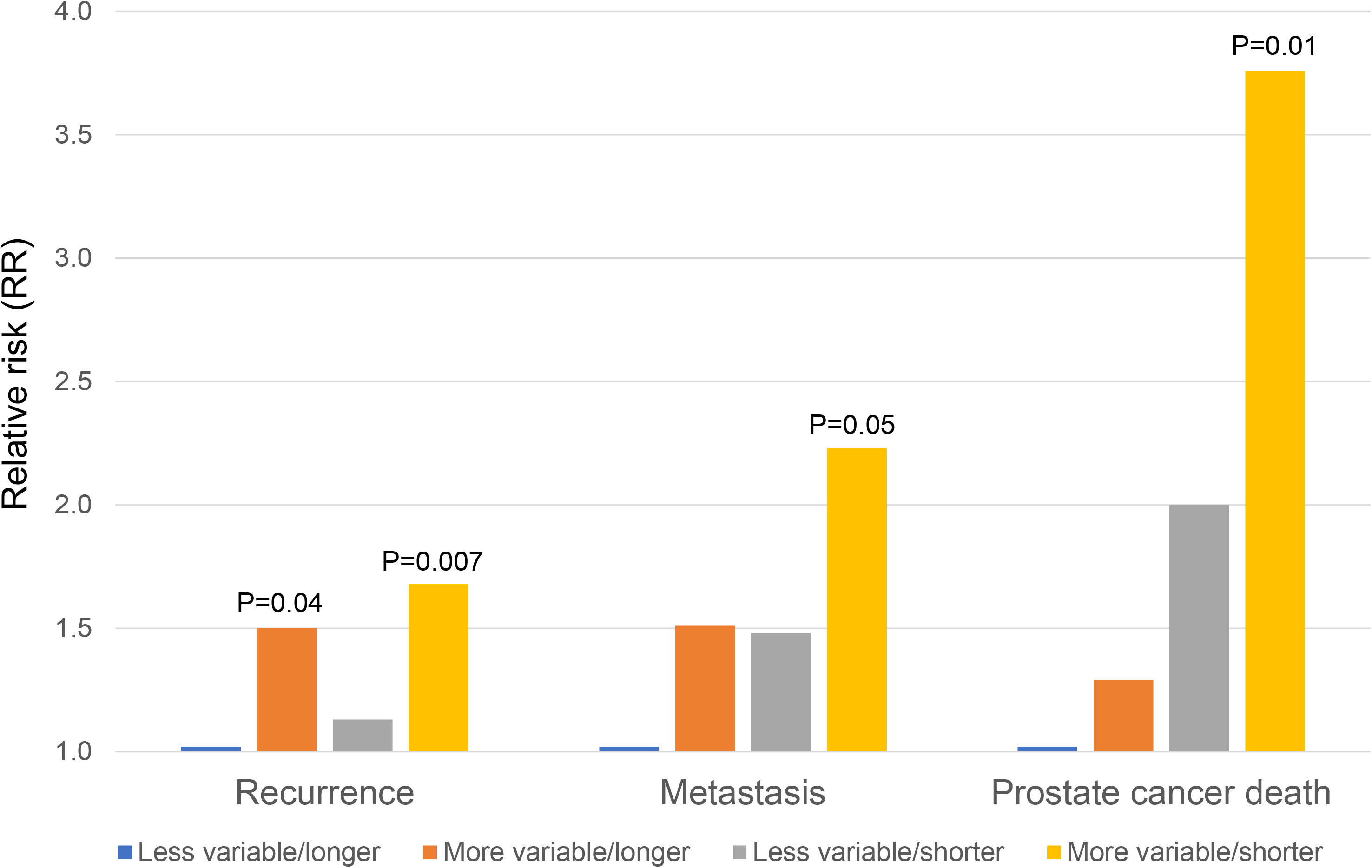
Meta-analytic summary associations between the telomere biomarker – combination of variability in telomere length among cancer cells and telomere length in cancer-associated stromal cells – and progression to recurrence, metastasis, and prostate cancer death after prostatectomy in five cohorts. Summary relative risks (RRs) from a meta-analysis of the HPFS, PHS, Johns Hopkins Recurrence, Johns Hopkins Intermediate-High Risk Study – I, and Johns Hopkins Intermediate-High Risk Study – II. Contributing RRs are adjusted for prognostic markers. P-value is for the comparison of the specified category with the less variable/longer combination of the telomere biomarker.

**Table 3.** Meta-analytic summary relative risks (RR) and 95% confidence intervals (CI) for the associations^1^ between the telomere biomarker – the combination of variability in telomere length among prostate cancer cells and telomere length in prostate cancer-associated stromal cells – and risk of recurrence, metastasis, and prostate cancer death after prostatectomy in 5 cohorts^2^

|  | Number of<br>contributing cohorts | Less variable/<br>longer | More variable/<br>longer | Less variable/<br>shorter | More variable/<br>shorter |
| --- | --- | --- | --- | --- | --- |
| Recurrence | 3 | 1.00<br>Reference | 1.50<br>1.03-2.18<br>P=0.04 | 1.13<br>0.81-1.56<br>P=0.47 | 1.68<br>1.16-2.44<br>P=0.007 |
| Metastasis | 4<sup>3</sup> | 1.00<br>Reference | 1.51<br>0.69-3.30<br>P=0.30 | 1.48<br>0.73-3.01<br>P=0.28 | 2.23<br>0.99-5.02<br>P=0.05 |
| Prostate cancer death | 2 | 1.00<br>Reference | 1.29<br>0.41-4.07<br>P=0.66 | 2.00<br>0.75-5.32<br>P=0.17 | 3.76<br>1.37-10.3<br>P=0.01 |

Bold denotes statistically significant at p<0.05 for a 2-sided test.
<sup>1</sup>Each contributing RR was adjusted for prognostic pathologic factors. RRs were summarized using inverse variance weights.
<sup>2</sup>50 prostate cancer deaths and 596 men were included in the original study in which we described the telomere biomarker.
<sup>3</sup>Includes all events (N=161) from the Johns Hopkins Intermediate-High Risk – II. When including only the 35 metastatic events, the number was too small distributed across the 4 telomere biomarker categories to obtain a stable estimate to meta-analyze.

**Supplement Table 8** shows study-specific RRs, and **Supplement Table 9** shows the summary HRs excluding the original HPFS data.

### The telomere biomarker and prostate cancer outcomes among men with intermediate disease

**Table 4** shows the summary associations between the telomere biomarker categories and prostate cancer outcomes when compared with the less variable/longer combination by Gleason sum (Grade Group). In men with Gleason 7 disease (Grade Groups 2/3), the more variable/shorter combination was statistically significantly associated with a higher risk of prostate cancer death (HR=9.18, 95% CI 1.14-74.0, P=0.037) when compared with the less variable/longer combination. All combinations of the telomere biomarker were statistically significantly associated with recurrence in men with Gleason sum >7 disease (Grade Groups 4/5) when compared with the less variable/longer combination.

**Table 4.** Meta-analytic summary RRs and 95% CIs for the associations^1^ between the telomere biomarker (combination of variability in telomere length among prostate cancer cells and telomere length in prostate cancer-associated stromal cells) and risk of recurrence, metastasis, and prostate cancer death after prostatectomy by Gleason sum category and PTEN status in 4 cohorts^2^

| Telomere biomarker | Gleason sum |  |  | PTEN status |  |
| --- | --- | --- | --- | --- | --- |
|  | <7<br>(grade group 1) | 7<br>(grade groups 2, 3) | >7<br>(grade groups 4, 5) | Intact | Null |
| **Recurrence** |  |  |  |  |  |
| Less variable/longer | 1.00<br>Reference | 1.00<br>Reference | 1.00<br>Reference | 1.00<br>Reference | 1.00<br>Reference |
| More variable/longer | 0.62<br>0.13-2.97 | 0.99<br>0.60-1.64 | 3.01<br>1.54-5.90<br>P=0.001 | 1.27<br>0.76-2.13 | 2.71<br>1.25-5.88<br>P=0.012 |
| Less variable/shorter | 0.16<br>0.04-0.73 | 0.86<br>0.55-1.32 | 2.00<br>1.05-3.82<br>P=0.035 | 0.89<br>0.57-1.40 | 1.59<br>0.78-3.24 |
| More variable/shorter | 1.94<br>0.64-5.93 | 1.25<br>0.74-2.11 | 2.63<br>1.28-5.37<br>P=0.008 | 1.40<br>0.83-2.39 | 3.05<br>1.37-6.78<br>P=0.006 |
| **Metastasis** |  |  |  |  |  |
| Less variable/longer | 1.00<br>Reference | 1.00<br>Reference | 1.00<br>Reference | 1.00<br>Reference | 1.00<br>Reference |
| More variable/longer | NE | 2.24<br>0.43-11.7 | 1.17<br>0.40-3.44 | 2.37<br>0.62-9.07 | 0.95<br>0.16-5.87 |
| Less variable/shorter | NE | 3.98<br>0.91-17.7 | 0.65<br>0.23-1.81 | 2.74<br>0.78-9.59 | 0.49<br>0.11-2.28 |
| More variable/shorter | NE | 4.27<br>0.81-22.4 | 1.49<br>0.54-4.14 | 3.85<br>0.99-14.9<br>P=0.051 | 2.72<br>0.25-30.1 |
| **Prostate cancer death** |  |  |  |  |  |
| Less variable/longer | 1.00<br>Reference | 1.00<br>Reference | 1.00<br>Reference | 1.00<br>Reference | 1.00<br>Reference |
| More variable/longer | NE | 0.23<br>0.01-4.06 | 1.31<br>0.36-4.76 | 0.87<br>0.14-5.56 | 3.99<br>0.28-55.8 |
| Less variable/shorter | NE | 4.16<br>0.54-31.9 | 0.88<br>0.27-2.84 | 1.70<br>0.36-8.01 | 2.89<br>0.27-36.8 |
| More variable/shorter | NE | 9.18<br>1.14-74.0<br>P=0.037 | 1.95<br>0.62-6.14 | 6.74<br>1.46-37.6<br>P=0.015 | 7.23<br>0.62-84.5 |

RR – Relative risk, CI – Confidence interval, NE – Not estimable (due to small sample size); Bold denotes statistically significant at $p < 0.05$ for a 2-sided test.
<sup>1</sup>Each contributing RR was adjusted for currently used prognostic pathologic factors. RRs were summarized using inverse variance weights.
<sup>2</sup>Of these, 50 prostate cancer deaths and 596 men were included in the original study in which we described the telomere biomarker. Of note, PTEN was not assessed in the Johns Hopkins Intermediate-High Risk Case-Cohort Study – II.

### The telomere biomarker, PTEN status, and prostate cancer outcomes

**Table 4** shows the summary associations between the telomere biomarker categories and prostate cancer outcomes when compared with the less variable/longer combination by PTEN status. In men with PTEN intact cancers, the more variable/shorter combination was positively associated with prostate cancer death (HR=6.74, 95% CI 1.46-37.6, P=0.015) and progression to metastasis (HR=3.85, 95% CI 0.99-14.9, P=0.051). In men with PTEN null cancers, telomere biomarker categories that included the more variable telomere length in cancer cell component were associated with a higher risk of recurrence than those with intact PTEN.

## DISCUSSION

In this study of 2,255 men across 5 cohorts, we confirmed that the telomere biomarker is associated with progression to metastasis and prostate cancer death in men surgically treated for clinically localized prostate cancer.

With the use of prostate specific antigen (PSA) screening in the US, most of the 248,530 prostate cancer cases (29) are detected when they are of small volume and apparently confined to the prostate, and thus should be curable by removal of the prostate. In the PSA era, despite having had their prostate removed, ∼25% experienced PSA re-elevation months to years later (30), with 25% of these occurring five or more years later (31). A third of men with PSA re-elevation developed overt metastases with a median time of 8 years after surgery (31) and 40% of these men died of their prostate cancer with a median time of 5 years after metastases are detected (31). However, following the 2012 and 2018 changes to the US Preventive Services Task Force prostate cancer screening recommendations, the stage at diagnosis has shifted toward more advanced disease in the US (32), which may result in a higher likelihood of recurrence among men treated by surgery. Thus, new molecular markers that improve prognostic accuracy are needed, particularly in men with intermediate risk disease.

In men with apparently organ-confined disease, the clinical tools currently used to predict disease behavior are inadequate and limit our ability to target men with optimal individualized treatment strategies. Furthermore, a thorough understanding of the molecular basis underlying aggressive cases is urgently needed for the continued development and refinement of prognostic indicators. We have now confirmed that the telomere biomarker is a promising molecular indicator of biological aggressiveness for use in prediction of prognosis. Telomere length variability and shortening are strongly associated with chromosomal instability, a hallmark of aggressive prostate cancer (33, 34).

In particular, we demonstrated that the more variable/shorter combination of the telomere biomarker is associated with prostate cancer death in men with Gleason 7 disease (Grade Groups 2/3). These men have the most variable clinical course, and are the group most in need of additional biomarkers for prognosis and treatment decision-making. We also showed that the more variable/shorter combination of the telomere biomarker is associated with both progression to metastasis and prostate cancer death in men with PTEN intact cancers. While PTEN null cancers have a worse prognosis, some men with PTEN intact cancers do progress. Thus, having an additional, independent biomarker may provide prognostic utility in this setting.

Only the more variable telomere length in cancer cells component of the telomere biomarker was associated with recurrence, as we previously observed in the HPFS (7). The exception was in men with Gleason sum >7 disease (>Grade Group 2 or 3), in whom each category of the telomere biomarker was associated with a 2 to 3 times increased risk of recurrence compared to the less variable/longer combination.

We previously discovered the telomere biomarker in the HPFS (7). In the current study, we followed the original 596 men for additional time and added 159 HPFS participants (represented across 2 TMAs) who were diagnosed and/or their tissue was arrayed subsequent to our initial study. This increased the number of prostate cancer deaths from 46 to 68 (48% increase) and the total follow-up time from 7,491 to 11,776 man-years (57% increase). While the patterns of association remained the same, the RRs were not as large as they were prior to these additions. For example, the RR of prostate cancer death for the more variable/shorter combination was 14.10 (95% CI 1.87-106) in our original study in the HPFS, 4.44 (95% CI 1.52- 13.0) in the expanded study in the HPFS, and 3.76 (95% CI 1.37-10.3) when combining across all 5 of the cohorts, remaining well above the null and statistically significant. While we do not know the explanation for why the RRs are smaller, a key difference is that in the original study we visually selected 30-50 of each cell type for image analysis. For the additional 2 HPFS TMAs as well as in the 4 other cohorts, we used a multiplexed set of markers for the image analysis software to identify the relevant cell types for inclusion and exclusion, and investigated all relevant cancer and stromal (excluding lymphocytes) cells that were in the image’s plane of focus. For the stromal cells, the new method identified hundreds to thousands of cells per cell type, likely capturing a different distribution of cells (compositionally and spatially) compared to the user-selected method. While the RR is smaller, with these additional HPFS and other cohort data provided greater precision (substantially narrower 95% CI).

Our study has a number of strengths. To confirm the telomere biomarker as prognostic for progression to metastatic disease and prostate cancer death, we used 5 studies of men who underwent prostatectomy developed with different criteria and different source populations (HPFS, PHS, Johns Hopkins). The cohorts we used had differing proportions of disease aggressiveness at diagnosis and likelihood of poor outcome (**Supplement Tables 1-5**). For example, the Johns Hopkins Recurrence Nested Case-Control Study was enriched for biochemical recurrence. In contrast, the two Johns Hopkins Intermediate-High Risk Case-Cohort Studies, were enriched for metastatic progression by design. Using these 5 studies allowed us to validate that the telomere biomarker is specific for progression to metastatic disease and prostate cancer death, but not recurrence, a finding that does not always progress to lethal prostate cancer.

We used a validated, optimized, semi-automated method of telomere length measurement that we developed (7) and semi-automated and optimized (25). The method allowed us to measure telomere length in fixed tissues, at single cell resolution in specific cell types, with all in-focus cells assessed. These attributes reduced or eliminated the possibility of systematic error that could be introduced by the operator’s bias in selecting individual cells for assessment, reduced or eliminated confounding by other cell types, and allowed for assessment of an important component of the biomarker – variability in telomere length among cancer cells (not the average length among these cells, as other methods would provide). Minimizing human error in measuring the components of the telomere biomarker is critical for routine clinical application of a molecular pathology-based prognostic tool.

Other aspects of the work warrant discussion. First, the method we developed does not determine actual telomere length (relative length is estimated and is linearly related to telomere length (28)) and does not determine chromosome-specific telomere length (chromosome-specific telomeric FISH probes for all chromosomes are not yet available). Second, the TMAs for each cohort were constructed using the largest cancer focus and/or with the highest Gleason pattern. We were not able to address whether the association between the telomere biomarker and poor outcome differs by which cancer foci was sampled. Nevertheless, we used the focus that is expected to impart the greatest risk. Third, we were not able to study recurrence, progression to metastasis, and prostate cancer death in each of the cohorts due to study designs and/or study populations used to construct the TMA sets we used. Fourth, given the 5 cohorts of men were surgically treated, we were not be able to address whether the telomere biomarker is associated with poor outcome in men undergoing radiation therapy, hormonal therapy, or other single or combined prostate cancer treatment modalities. Fifth, our goal is to develop a tool for use at the time of prostatectomy to aid in decision-making about the need for additional treatment and more intensive surveillance. A complementary research question is whether the telomere biomarker has prognostic utility at the time of biopsy prior to any treatment. To determine whether men classified as low risk by the telomere biomarker might not require treatment at all, optimally, we would study men who are confirmed to have prostate cancer by biopsy and who are not treated and followed for outcome (i.e. men enrolled in active surveillance). However, men selected for active surveillance are, by definition, at a low risk of a poor outcome, and thus a very large study with long follow-up would be required to test the prognostic utility of the telomere biomarker.

In conclusion, we documented the robustness of the telomere biomarker as a prognostic tool for lethal prostate cancer. We focused on the length of the telomeres for this biomarker because abnormally shortened telomeres are intimately involved in promoting carcinogenesis, including by promoting the accumulation of chromosomal instability, a hallmark of prostate cancer (35). We demonstrated that the telomere biomarker captures information in the prostatectomy specimen about tumor behavior beyond currently used indicators, thereby identifying men who are more or less likely to benefit from additional treatment. Thus, we expect that the telomere biomarker could be used to stratify men for individualized therapeutic strategies and has the potential of increasing the benefit to risk ratio for men and reducing healthcare costs associated with prostate cancer.

## Data Availability

All data produced in the present work are contained in the manuscript

## Acknowledgments

The Prostate Cancer Biorepository Network (PCBN) provided some TMAs for this study, which is supported by the Department of Defense Prostate Cancer Research Program Award (W81XWH-15-2-0062, W81XWH-18-2-0015). We could like to thank the following state cancer registries for their help: AL, AZ, AR, CA, CO, CT, DE, FL, GA, ID, IL, IN, IA, KY, LA, ME, MD, MA, MI, NE, NH, NJ, NY, NC, NC, OH, OK, OR, PA, RI, SC, TN, TX, VA, WA, WY. We are grateful to the participants and staff of the Health Professionals Follow-up Study. The authors assume full responsibility for the analyses and interpretation of these data.

## SUPPLEMENTAL METHODS

### Health Professionals Follow-up Study (HPFS)

The HPFS is an ongoing prospective cohort study on risk factors for cancer and other chronic diseases (https://www.hsph.harvard.edu/hpfs). In 1986, 51,529 men of ages 40 to 75 years were enrolled. On each follow-up questionnaire, we asked the men to report a diagnosis of prostate cancer, which are confirmed by medical records and pathology reports (for 94.5%). Tumor–node–metastasis (TNM) stage and PSA concentration at diagnosis were abstracted from these records. The men were followed from the date of their surgery to 2014. Diagnosis of recurrence and progression to distant metastasis (to bone or other organs) was collected by mailed questionnaire and then confirmed by the treating doctor. Investigators learned of a participant’s death from family members, the postal system, or by searches of the National Death Index. Men were classified as having died from their prostate cancer (underlying cause on the death certificate) if they also had documented extensive metastatic disease.

### Physicians’ Health Study

The parent study, Physicians’ Health Study (PHS) was a large, randomized chemoprevention trials for cancer and cardiovascular disease (1). The PHS included 29,067 male physicians who were 40 to 84 years old and who did not have a history of cancer at the time of randomization in 1983. Prostate cancer diagnoses were identified by self-report on a mailed questionnaire and were confirmed by medical record and pathology report review. Data abstracted from these records included Gleason sum, TNM staging, and PSA concentration at the time of diagnosis. The men were followed from the date of their surgery to 2014. Bony metastases were confirmed by the treating physician. Deaths were ascertained by a search of the National Death Index or by reports from the US Postal Service or next of kin. Men were classified as dying from their prostate cancer if they had evidence of extensive metastatic disease; the Endpoint Committee made this determination. Mortality follow-up is >99% complete for this cohort.

### Telomere-specific FISH and Immunostaining

The staining methods are adapted from previously described studies (2–4), with the following modifications. Deparaffinized TMA slides were hydrated through a graded ethanol series, placed in deionized water, followed by deionized water plus 0.1% Tween-20. The TMA slides were steamed for 25 minutes in citrate buffer (catalog # H-3300; Vector Laboratories), removed, and allowed to cool at room temperature for 10 minutes. The TMA slides were washed in deionized water, dehydrated through a graded ethanol series, and then air-dried. Thirty-five μL of the telomere-specific peptide nucleic acid (PNA) probe [0.33 μg/mL PNA (CCCTAACCCTAACCCTAA with the N-terminal covalently linked to Cy3; Panagene) in 70% formamide and 10 mmol/L Tris, pH 7.5)] was applied, coverslipped, and denatured by incubation for 5 minutes at 84°C. The TMA slides were then hybridized overnight in the dark in a humidified hybridization chamber. Next, the slides were washed in PNA wash buffer (70% formamide, 29% deionized distilled water; 1% 1 M Tris-Cl, pH 7.5) and then in PBST. Slides were incubated for 30 min at room temperature with serum-free protein block (DAKO; cat# X0909), washed in PBST, and then incubated for 2 hrs at room temperature with the following antibody cocktail diluted in antibody dilution buffer (Ventana; cat# ADB250): basal-specific anti-cytokeratin primary antibody (34BE12, Enzo; 1:50 dilution), an anti-NKX3.1 primary antibody (Athena; 1:1000 dilution), an anti-FOXA1 primary antibody (Abcam; 1:500 dilution), an anti-CD3 primary antibody (DAKO; 1:200 dilution), and an anti-CD20 primary antibody (Abcam; 1:20 dilution). After incubation, the slides were washed in PBST and incubated for 30 min at room temperature with anti-rabbit IgG fraction Alexa Fluor 488 and anti-mouse IgG fraction Alexa Fluor 647 (secondary antibodies) in PBS at a 1:100 dilution. Following washes in PBST and deionized water, slides were stained with 4′-6-diamidino-2-phenylindole (DAPI) solution (500 ng/ml in deionized distilled water) for 10 min. The TMA slides were then mounted with Prolong antifade mounting medium (catalog no.: P-7481; Molecular Probes)

---

**Supplement Table 1.**
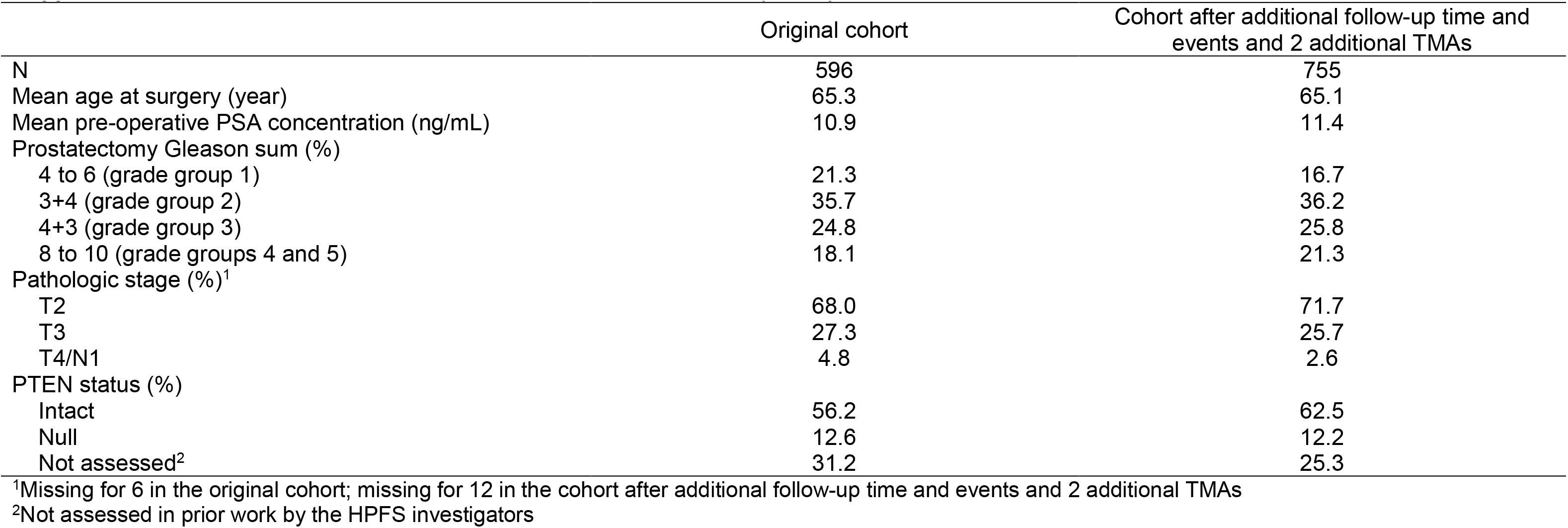
Characteristics of the Health Professionals Follow-up Study

|  | Original cohort | Cohort after additional follow-up time and events and 2 additional TMAs |
| --- | --- | --- |
| N | 596 | 755 |
| Mean age at surgery (year) | 65.3 | 65.1 |
| Mean pre-operative PSA concentration (ng/mL) | 10.9 | 11.4 |
| Prostatectomy Gleason sum (%) |  |  |
| 4 to 6 (grade group 1) | 21.3 | 16.7 |
| 3+4 (grade group 2) | 35.7 | 36.2 |
| 4+3 (grade group 3) | 24.8 | 25.8 |
| 8 to 10 (grade groups 4 and 5) | 18.1 | 21.3 |
| Pathologic stage (%)<sup>1</sup> |  |  |
| T2 | 68.0 | 71.7 |
| T3 | 27.3 | 25.7 |
| T4/N1 | 4.8 | 2.6 |
| PTEN status (%) |  |  |
| Intact | 56.2 | 62.5 |
| Null | 12.6 | 12.2 |
| Not assessed<sup>2</sup> | 31.2 | 25.3 |

<sup>1</sup>Missing for 6 in the original cohort; missing for 12 in the cohort after additional follow-up time and events and 2 additional TMAs
<sup>2</sup>Not assessed in prior work by the HPFS investigators

**Supplement Table 2.** Characteristics of the Physicians’ Health Study

|  | Cohort spotted on TMAs | Cohort with useable TMA spots<sup>1</sup> |
| --- | --- | --- |
| N | 500 | 151 |
| Mean age at surgery (year) | 65.1 | 66.0 |
| Mean pre-operative PSA concentration (ng/mL) | 12.8 | 12.1 |
| Prostatectomy Gleason sum (%) |  |  |
| 4 to 6 (grade group 1) | 44 | 22 |
| 7 (grade groups 2 and 3) | 41 | 53 |
| 8 to 10 (grade groups 4 and 5) | 15 | 22 |
| Pathologic stage (%) |  |  |
| T2 | 72 | 74 |
| T3 | 24 | 21 |
| T4/N1 | 4 | 5 |
| PTEN status (%) |  |  |
| Intact | 38.2 | 42.4 |
| Null | 11.0 | 13.2 |
| Not assessed | 50.8 | 44.4 |

<sup>1</sup>Reasons that spots were not useable included some the spots no longer contained tumor (due to the repeated sectioning for multiple prior studies), the tissue fell out of the section matrix, or was missing initially, had high auto-fluorescent background (depending on the TMA, ~15-20% of spots) that prohibited determination of telomere length.

**Supplement Table 3.** Characteristics of 376 prostate cancer biochemical recurrence cases and 376 matched controls in the Johns Hopkins Recurrence Nested Case-Control Study^1^

|  | Cases | Controls | P |
| --- | --- | --- | --- |
| Mean age at surgery (year) ± standard deviation | 58.9 ± 6.2 | 59.0 ± 5.9 | Matched |
| Race (%) |  |  |  |
| White | 86.4 | 89.6 |  |
| Black | 8.8 | 7.2 | Matched |
| Other race/ethnicity | 4.8 | 3.2 |  |
| Follow-up time (yr), median, IQR | 2 (1-3) | 6 (3-8) | <0.0001 |
| Pre-operative PSA concentration (ng/mL), median (IQR) | 12.2 (6.0-14.3) | 8.8 (6.1-14.3) | 0.1 |
| Prostatectomy Gleason sum (%) |  |  |  |
| ≤ 6 (grade group 1) | 15.1 | 15.4 |  |
| 3+4 (grade group 2) | 36.7 | 47.6 | Matched |
| 4+3 (grade group 3) | 21.8 | 12.8 |  |
| >7 (grade groups 4 and 5) | 26.3 | 24.2 |  |
| Pathologic stage (%) |  |  |  |
| T2 | 12.8 | 13.0 |  |
| T3a | 10.1 | 17.6 | Matched |
| T3b or N1 | 77.1 | 69.4 |  |
| Positive surgical margins (%) | 34.8 | 24.7 | 0.001 |
| PTEN status (%) |  |  |  |
| Intact | 47.9 | 57.5 | 0.001 |
| Null | 39.6 | 37.0 |  |
| Not assessed | 12.5 | 5.6 |  |

<sup>1</sup>Controls were sampled using incidence density sampling; the number of unique men is 549.
<sup>2</sup>IQR – Interquartile range

**Supplement Table 4.** Characteristics of men in the Johns Hopkins Intermediate-High Risk Case-Cohort Study - I^1^

|  | All men in sample | Subcohort | Cases outside of the subcohort |
| --- | --- | --- | --- |
|  | 306 | 253<sup>2</sup> | 53 |
| Median age at surgery, year (IQR<sup>3</sup>) | 60 (56-64) | 60 (56-64) | 59 (56-64) |
| White (%) | 90.2 | 89.3 | 94.3 |
| Prostatectomy Gleason sum (%) |  |  |  |
| ≤7 (grade groups 1, 2, and 3) | 55.6 | 62.4 | 22.6 |
| 8 (grade group 4) | 11.8 | 11.5 | 13.2 |
| 9 (grade group 5) | 31.7 | 24.9 | 64.2 |
| Missing | 1.0 | 1.2 | 0 |
| Pathologic stage (%) |  |  |  |
| T2 | 24.5 | 28.5 | 5.7 |
| T3a | 47.1 | 49.4 | 35.8 |
| T3b | 27.5 | 20.9 | 58.5 |
| Missing | 1.0 | 1.2 | 0 |
| Positive surgical margins (%) | 27.5 | 24.9 | 39.6 |
| Median preoperative PSA concentration, ng/mL (IQR<sup>3</sup>) | 10.2 (6.4-15.1) | 10.1 (6.3-15.0) | 10.8 (6.7-15.1) |
| PTEN status (%) |  |  |  |
| Intact | 63.7 | 67.2 | 47.2 |
| Null | 36.3 | 32.8 | 52.8 |

<sup>1</sup>With useable TMA spots.<sup>2</sup>Includes 62 men who later become cases.<sup>3</sup>IQR – interquartile range

**Supplement Table 5.**
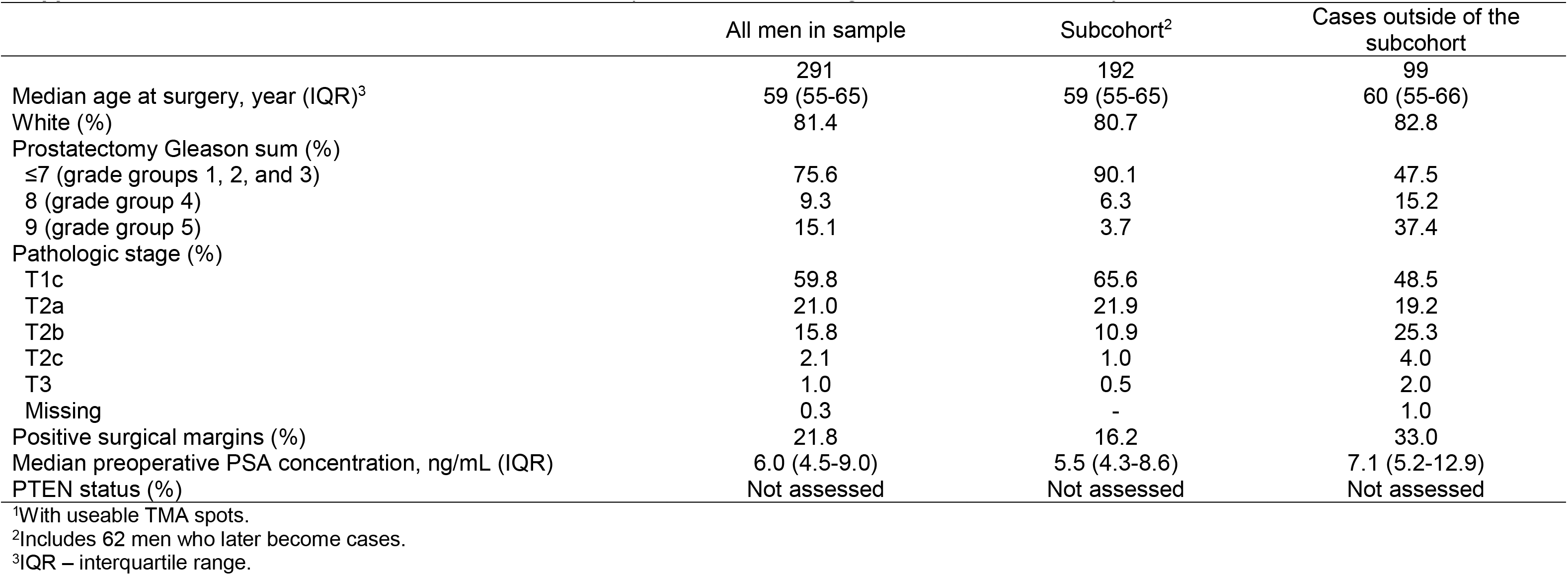
Characteristics of men in the Johns Hopkins Intermediate-High Risk Case-Cohort Study – II^1^

**Supplement Table 6.** Associations1 of more variable telomere length among prostate cancer cells and shorter telomere length in prostate cancer-associated stromal cells with risk of recurrence, metastasis, and prostate cancer death after prostatectomy in 5 cohorts

|  |  |  |  |  |  |  |  |  |  |  |  |  |  |  |  |  |  |  |  |  |
| --- | --- | --- | --- | --- | --- | --- | --- | --- | --- | --- | --- | --- | --- | --- | --- | --- | --- | --- | --- | --- |
| Prostate cancer outcome | HPFS<sup>2</sup> |  |  |  | PHS<sup>2</sup> |  |  |  | Johns Hopkins Recurrence<sup>3</sup> |  |  |  | Johns Hopkins Intermediate-High Risk - I<sup>4</sup> |  |  |  | Johns Hopkins Intermediate-High Risk – II<sup>4</sup> |  |  |  |
|  | N | Person-years | More variable length among cancer cells | Shorter length in stromal cells | N | Person-years | More variable length among cancer cells | Shorter length in stromal cells | Cases | Controls | More variable length among cancer cells | Shorter length in stromal cells | Cases | Sub-cohort | More variable length among cancer cells | Shorter length in stromal cells | Cases | Sub-cohort | More variable length among cancer cells | Shorter length in stromal cells |
| Recurrence | 747 | 9102 | 1.28<br>0.97-1.68 | 1.05<br>0.79-1.39 | 149 | 1668 | 1.35<br>0.74-2.47 | 0.57<br>0.31-1.03 | 376 | 376 | 1.82<br>1.18-2.83 | 0.95<br>0.62-1.45 | - | - | - | - | - | - | - | - |
| Metastasis | 747 | 11291 | 1.23<br>0.59-2.60 | 1.26<br>0.57-2.79 | 149 | 2067 | 10.93<br>0.40-298 | 0.15<br>0.01-2.01 | - | - | - | - | 115 | 253 | 1.26<br>0.76-2.07 | 1.18<br>0.64-2.18 | 161 | 192 | 1.38<sup>5</sup><br>0.68-2.81 | 1.23<sup>5</sup><br>0.62-2.43 |
| Prostate cancer death | 755 | 12305 | 1.29<br>0.78-2.15 | 2.55<br>1.36-4.78 | 151 | 2092 | 2.02<br>0.55-7.46 | 0.58<br>0.18-1.91 | - | - | - | - | - | - | - | - | - | - | - | - |

<sup>1</sup>Of these, 50 prostate cancer deaths and 596 men were included in the original study in which we described the telomere biomarker. HR and 95% CI estimated from a Cox proportional hazards regression model and adjusted for age, year of surgery, prostatectomy Gleason sum, pathologic stage, pre-operative serum PSA.
<sup>2</sup>HR and 95% CI estimated from a Cox proportional hazards regression model and adjusted for age, year of surgery, prostatectomy Gleason sum, pathologic stage, and pre-operative serum PSA concentration.
<sup>3</sup>OR and 95% CI estimated from a conditional logistic regression model adjusted for year of surgery, primary and secondary Gleason pattern, pre-operative PSA, and surgical margins (cases and controls were matched on age, race, prostatectomy Gleason sum, and pathologic stage).
<sup>4</sup>HR and 95% CI estimated from a Cox proportional hazards regression model using robust variance correction and adjusted for age, race, pathologic stage, prostatectomy Gleason sum, year of surgery, pre-operative PSA, and surgical margins.
<sup>5</sup>Outcome is the combination of rapidly rising PSA, which is indicative of likely progression to metastasis, and metastases; restricting to metastasis only (N=35): more variable length among cancer cells HR=3.58, 95% CI 0.78-16.33, shorter length among stromal cells HR=1.00, 95% CI 0.23-4.34.

**Supplement Table 7.**
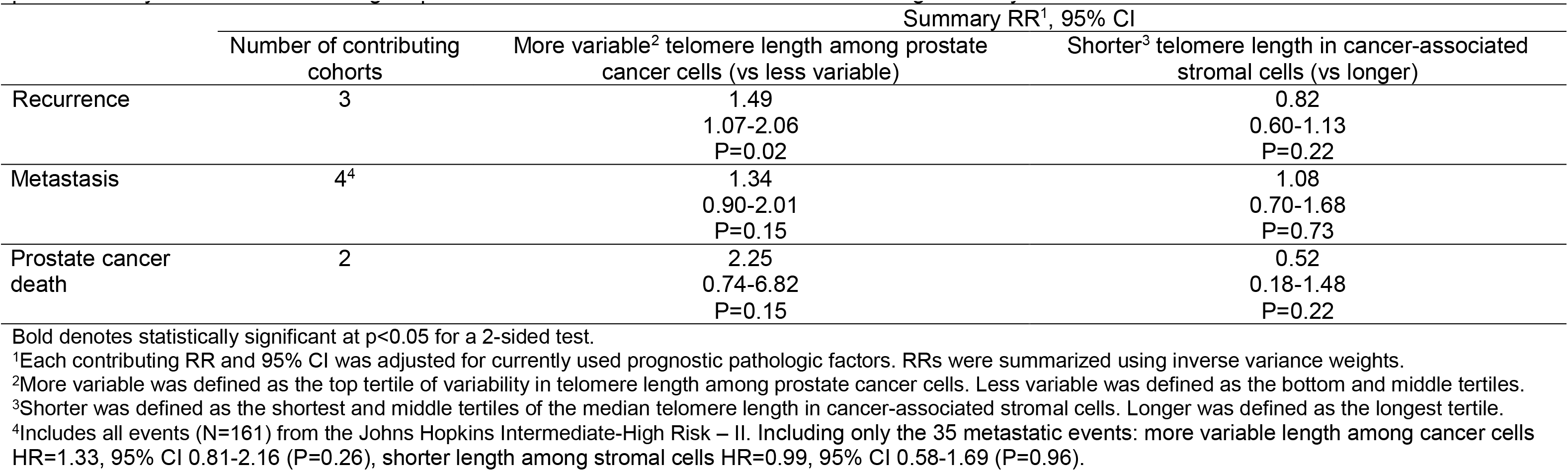
Meta-analytic summary relative risks (RR) and 95% confidence intervals (CI) of more variable telomere length among prostate cancer cells and shorter telomere length in prostate cancer-associated stromal cells with risk of recurrence, metastasis, and prostate cancer death after prostatectomy in 5 cohorts excluding 50 prostate cancer deaths and 596 men from the original study in which we described the telomere biomarker

|  | Number of contributing cohorts | Summary RR<sup>1</sup>, 95% CI |  |
| --- | --- | --- | --- |
|  |  | More variable<sup>2</sup> telomere length among prostate cancer cells (vs less variable) | Shorter<sup>3</sup> telomere length in cancer-associated stromal cells (vs longer) |
| Recurrence | 3 | 1.49<br>1.07-2.06<br>P=0.02 | 0.82<br>0.60-1.13<br>P=0.22 |
| Metastasis | 4<sup>4</sup> | 1.34<br>0.90-2.01<br>P=0.15 | 1.08<br>0.70-1.68<br>P=0.73 |
| Prostate cancer death | 2 | 2.25<br>0.74-6.82<br>P=0.15 | 0.52<br>0.18-1.48<br>P=0.22 |

Bold denotes statistically significant at p<0.05 for a 2-sided test.
<sup>1</sup>Each contributing RR and 95% CI was adjusted for currently used prognostic pathologic factors. RRs were summarized using inverse variance weights.
<sup>2</sup>More variable was defined as the top tertile of variability in telomere length among prostate cancer cells. Less variable was defined as the bottom and middle tertiles.
<sup>3</sup>Shorter was defined as the shortest and middle tertiles of the median telomere length in cancer-associated stromal cells. Longer was defined as the longest tertile.
<sup>4</sup>Includes all events (N=161) from the Johns Hopkins Intermediate-High Risk – II. Including only the 35 metastatic events: more variable length among cancer cells HR=1.33, 95% CI 0.81-2.16 (P=0.26), shorter length among stromal cells HR=0.99, 95% CI 0.58-1.69 (P=0.96).

**Supplement Table 8.** Associations between the telomere biomarker – the combination of variability in telomere length among cancer cells and telomere length in cancer-associated stromal cells – and risk of recurrence, metastasis, and prostate cancer death after prostatectomy in 5 cohorts

| Telomere biomarker<sup>1</sup> | HPFS Prostate Cancer Study<sup>1,2</sup> |  |  | PHS<sup>2</sup> |  |  | Johns Hopkins Recurrence<sup>3</sup> |  |  | Johns Hopkins Intermediate-High Risk - I<sup>4</sup> |  |  | Johns Hopkins Intermediate-High Risk – II<sup>4,5</sup> |  |  |
| --- | --- | --- | --- | --- | --- | --- | --- | --- | --- | --- | --- | --- | --- | --- | --- |
|  | No. | PY | HR, 95% CI | No. | PY | HR, 95% CI | Cases | Controls | OR, 95% CI | Cases | Subcohort | HR, 95% CI | Cases | Subcohort | HR, 95% CI |
| **Recurrence** |  |  |  |  |  |  |  |  |  |  |  |  |  |  |  |
| Less variable/longer | 31 | 1502 | 1.00<br>Reference | 6 | 217 | 1.00<br>Reference | 43 | 50 | 1.00<br>Reference | - | - | - | - | - | - |
| More variable/longer | 40 | 1318 | 1.18<br>0.73-1.90 | 13 | 357 | 2.21<br>0.76-6.45 | 115 | 76 | 2.21<br>1.06-4.62 | - | - | - | - | - | - |
| Less variable/shorter | 103 | 4620 | 1.05<br>0.70-1.58 | 23 | 874 | 1.07<br>0.41-2.80 | 157 | 202 | 1.38<br>0.71-2.64 | - | - | - | - | - | - |
| More variable/shorter | 48 | 1229 | 1.48<br>0.94-2.34 | 9 | 219 | 1.51<br>0.52-4.37 | 61 | 48 | 2.65<br>1.17-5.97 | - | - | - | - | - | - |
| **Metastasis** |  |  |  |  |  |  |  |  |  |  |  |  |  |  |  |
| Less variable/longer | 3 | 1842 | 1.00<br>Reference | 1 | 280 | NE | - | - | - | 15 | 29 | 1.00<br>Reference | 6 | 22 | 1.00<br>Reference |
| More variable/longer | 6 | 1729 | 1.69<br>0.42-6.89 | 2 | 475 | NE | - | - | - | 17 | 41 | 1.17<br>0.38-3.55 | 30 | 41 | 2.41<br>0.41-14.3 |
| Less variable/shorter | 15 | 5582 | 1.64<br>0.47-5.70 | 0 | 1018 | NE | - | - | - | 64 | 99 | 1.20<br>0.44-3.25 | 72 | 93 | 2.26<br>0.40-12.7 |
| More variable/shorter | 6 | 1642 | 1.98<br>0.49-8.02 | 2 | 294 | NE | - | - | - | 19 | 22 | 1.87<br>0.62-5.59 | 17 | 10 | 7.62<br>0.66-88.1 |
| **Prostate cancer death** |  |  |  |  |  |  |  |  |  |  |  |  |  |  |  |
| Less variable/longer | 4 | 1982 | 1.00<br>Reference | 1 | 282 | 1.00<br>Reference | - | - | - | - | - | - | - | - | - |
| More variable/longer | 7 | 1898 | 1.19<br>0.34-4.17 | 3 | 483 | 1.96<br>0.11-33.9 | - | - | - | - | - | - | - | - | - |
| Less variable/shorter | 32 | 6084 | 2.50<br>0.88-7.10 | 11 | 1029 | 0.38<br>0.02-6.47 | - | - | - | - | - | - | - | - | - |
| More variable/shorter | 24 | 1812 | 4.44<br>1.52-13.0 | 2 | 298 | 1.04<br>0.05-20.6 | - | - | - | - | - | - | - | - | - |

NE – Not estimable, Bold denotes statistically significant at p<0.05 for a 2-sided test.
<sup>1</sup>Of these, 50 prostate cancer deaths and 596 men were included in the original study in which we described the telomere biomarker. Adjusted for age, year of surgery, prostatectomy Gleason sum, pathologic stage, pre-operative serum PSA.
<sup>2</sup>Adjusted for age, year of surgery, prostatectomy Gleason sum, pathologic stage, and pre-operative serum PSA concentration.
<sup>3</sup>Adjusted for year of surgery, primary and secondary Gleason pattern, pre-operative PSA, and surgical margins (cases and controls were matched on age, race, prostatectomy Gleason sum, and pathologic stage).
<sup>4</sup>Adjusted for age, race, pathologic stage, prostatectomy Gleason sum, year of surgery, pre-operative PSA, and surgical margins.
<sup>5</sup>Outcome is the combination of rapidly rising PSA, which is indicative of likely progression to metastasis, and metastases. When including only the 35 metastatic events, the number was too small distributed across the 4 telomere biomarker categories to obtain a stable estimates.

**Supplement Table 9.** Meta-analytic summary RRs and 95% CIs for the associations^1^ between the telomere biomarker (combination of variability in telomere length among prostate cancer cells and telomere length in prostate cancer-associated stromal cells) and risk of recurrence, metastasis, and prostate cancer death after prostatectomy in the 5 cohorts excluding 50 prostate cancer deaths and 596 men from the original study in which we described the telomere biomarker

|  | Number of<br>contributing cohorts | Less variable/ longer | More variable/ longer | Less variable/ shorter | More variable/ shorter |
| --- | --- | --- | --- | --- | --- |
| Recurrence | 3 | 1.00<br>Reference | 1.57<br>0.91-2.73<br>P=0.11 | 1.01<br>0.63-1.63<br>P=0.97 | 1.70<br>0.96-3.01<br>P=0.07 |
| Metastasis | 4<sup>2</sup> | 1.00<br>Reference | 1.43<br>0.56-3.68<br>P=0.46 | 1.24<br>0.53-2.89<br>P=0.62 | 2.11<br>0.82-5.41<br>P=0.12 |
| Prostate cancer death | 2 | 1.00<br>Reference | 1.02<br>0.14-7.55<br>P=0.93 | 0.21<br>0.02-1.78<br>P=0.15 | 0.88<br>0.12-6.70<br>P=0.90 |

Bold denotes statistically significant at p<0.05 for a 2-sided test.
<sup>1</sup>Each contributing RR was adjusted for prognostic pathologic factors. RRs were summarized using inverse variance weights.
<sup>2</sup>Includes all events (N=161) from the Johns Hopkins Intermediate-High Risk – II. When including only metastatic events (N=35), associations were not estimable.

